# A STUDY OF ETIOLOGICAL AND CLINICAL PROFILE OF PATIENTS WITH LMN CRANIAL NERVE PALSY IN A TERTIARY CARE CENTER AT THANJAVUR MEDICAL COLLEGE AND HOSPITAL

**DOI:** 10.1101/2021.03.06.21253049

**Authors:** B. Nandini Priyanka, A. Gunasekaran, M. Thangaraj

## Abstract

**Background:** Neurological presentation with isolated/multiple cranial nerve palsy is normal and its different causes incorporate infection, autoimmune, neoplastic, and inflammatory pathologies. The etiological range may rely on geological areas. We attempted this study to investigate the clinical range and etiological profile of numerous cranial nerve palsy.

**Materials and methods:** This planned observational study was led from January 2020 to January 2021. All the patients with cranial nerve palsy coming to the neurology OPD were taken for examinations. Essential targets were to characterize anatomical disorder/cranial nerve combinations and to set up etiology. The primary goals were to examine related components. Patients with neuromuscular junction disorder, anterior horn cell disease, myopathies, first and second cranial nerve dysfunction were excluded from the study. All patients went through an organized convention of clinical assessment, examinations, and few particular examinations as per clinical protocol for analysis.

**Results:** Cavernous sinus was the commonest anatomical condition of different cranial nerve paralyses and tuberculous disease was the commonest cause in this investigation.7^th^ cranial nerve was the common isolated nerve involved with idiopathic ethology, diabetes was the most common cause overall found with third nerve involvement.

## BACKGROUND

Isolated single/more than one LMN cranial neuropathies or disorder is a commonly encountered medical problem. The assessment of these patients is often overwhelming due to an extensive range of etiologies in addition to the capability for devastating neurologic consequences. Dysfunction of the cranial nerves can arise because of the lesion anywhere of their route from the intrinsic brainstem to their peripheral courses [1].

The afferent and efferent connections of the cranial nerves traverse through the meninges, subarachnoid space, bony structures of the skull, and superficial smooth tissues. The cranial nerve nuclei lie within the brain stem, as a result, the intra-axial pathologic system may additionally present, to begin with, the most effective cranial nerve disorder too. Therefore, many such pathologic approaches are manifested by using cranial nerve disorder [2]. There can be involvement of homologous nerves on the two aspects (i.e., bilateral facial palsy) or one-of-a-kind nerves on the identical or contra-lateral aspect. In some conditions, a set of nerves is involved in a discrete anatomic vicinity constituting distinct anatomical syndrome.

Maximum of the literature regarding etiologies of single/more than one cranial neuropathies include case reviews or case series. The biggest stated retrospective collection by way of Keane had 979 instances, collected over 12 months period [3]. There had been diverse locations and reasons for cranial nerve involvement and Tumours were the most common reason, however, an extensive proportion remained idiopathic.[3] vast and sequential involvements of cranial nerves point in the direction of the opportunity of malignant infiltration of meninges, however, confirmation of diagnosis might not be feasible without biopsy or earlier than post-mortem [4].

In the Indian subcontinent, where infectious diseases are predominant, tuberculous meningitis is an important motive of cranial nerve palsies that is seen in nearly one-third of cases. The presence of cranial neuropathy is also associated with poor final results [5]. Considering that there may be a paucity of Indian studies at the etiological spectrum of LMN cranial nerve palsy, we determined to adopt this observation with the purpose to evaluate clinical spectrum and causes of LMN cranial nerve palsies in tertiary healthcare institutions of India.

## PREVALENCE

### STUDY MATERIALS

The principal investigator should include details of the following

#### A. Study design

Cross-sectional observational

#### B. Study setting

In-Patients of Neurology Department At Government Thanjavur Medical College and Hospital

#### C. Study subjects

Patients who are Attending Neurology opd with Symptoms and Signs of Cranial Neuropathies will be taken as Subjects

#### D. Inclusion criteria

All the consecutive patients of single/multiple lower motor neuron cranial palsies presenting to the neurology department were included in the studies.

#### E. Exclusion criteria

First and Second Cranial Nerve Involvement, NMJ Disorders, And Muscle Disorders are Excluded.

#### F. Sample size

For observational studies, the sample size is justified.

#### G. Period Of Study

January 2020 to January 2021

#### H. Methods

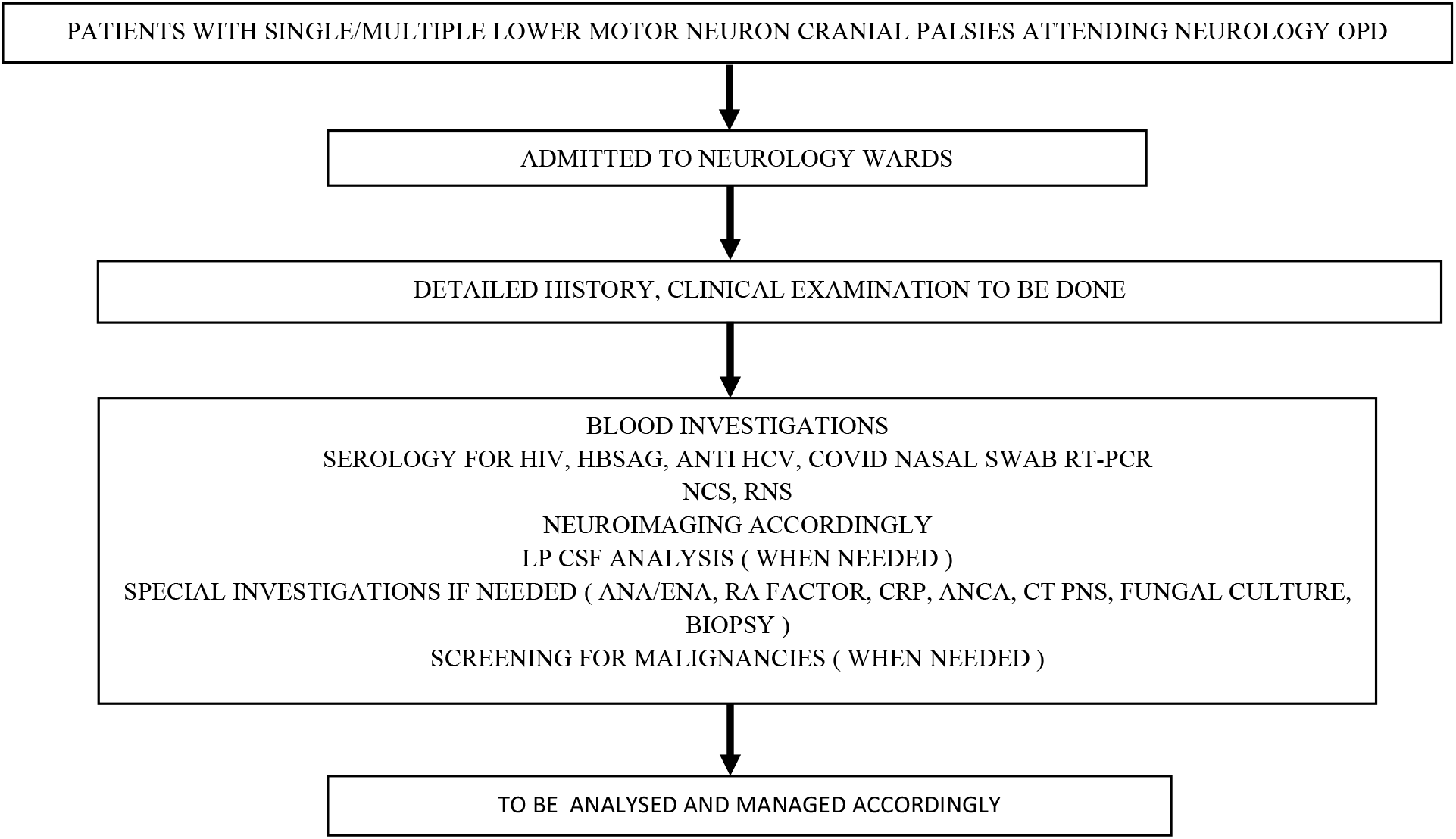

#### I. Parameter analyzed

Age, sex, onset, duration, progression, side of involvement, number of cranial nerves involved, blood routine investigations, Diabetic status, Hypertension status, Nerve conduction study, MRI imaging

#### J. Statistical analysis

a. Chi-square test – to compare the frequency/proportions between the groups with sample >30
b. Pearson and spearman’s correlation test – to find the strength of association between two parameters of the parametric and non-parametric distribution, respectively.

## RESULTS

### 1. GENERAL RESULTS

#### A. Sex, Age Wise And Cranial Nerve Distribution

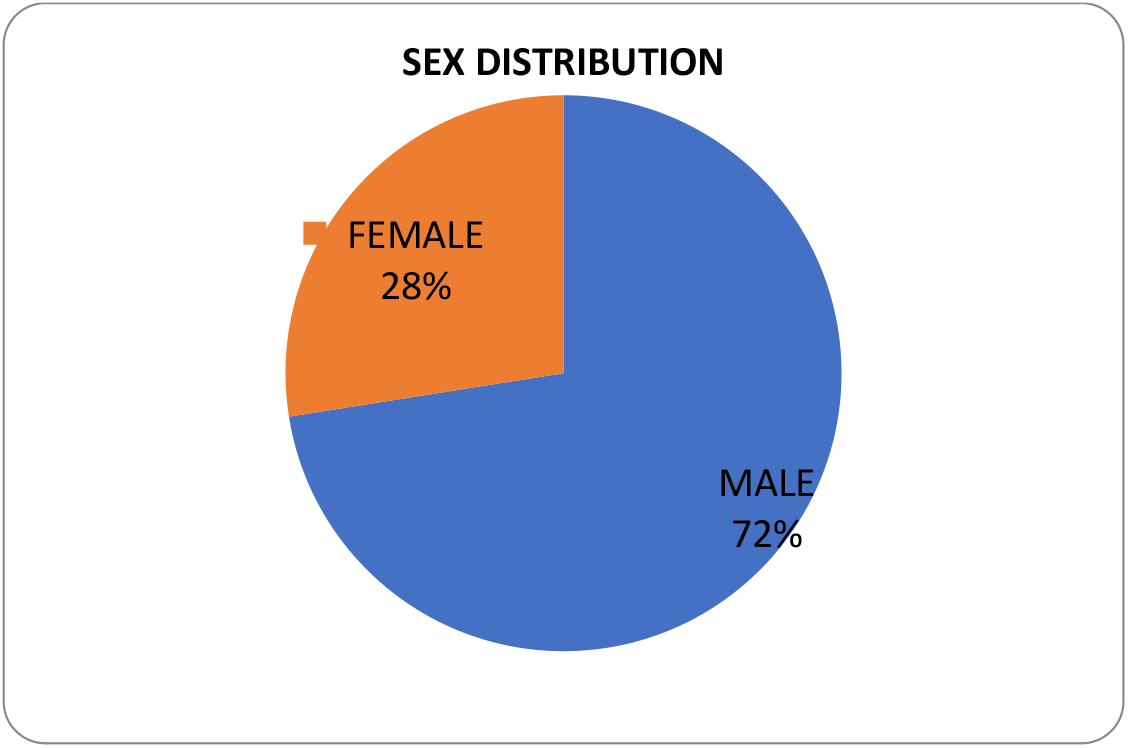

Overall, out of 80 patients, 72% were males(58), 28% were females (22); the Most common gender population were males.

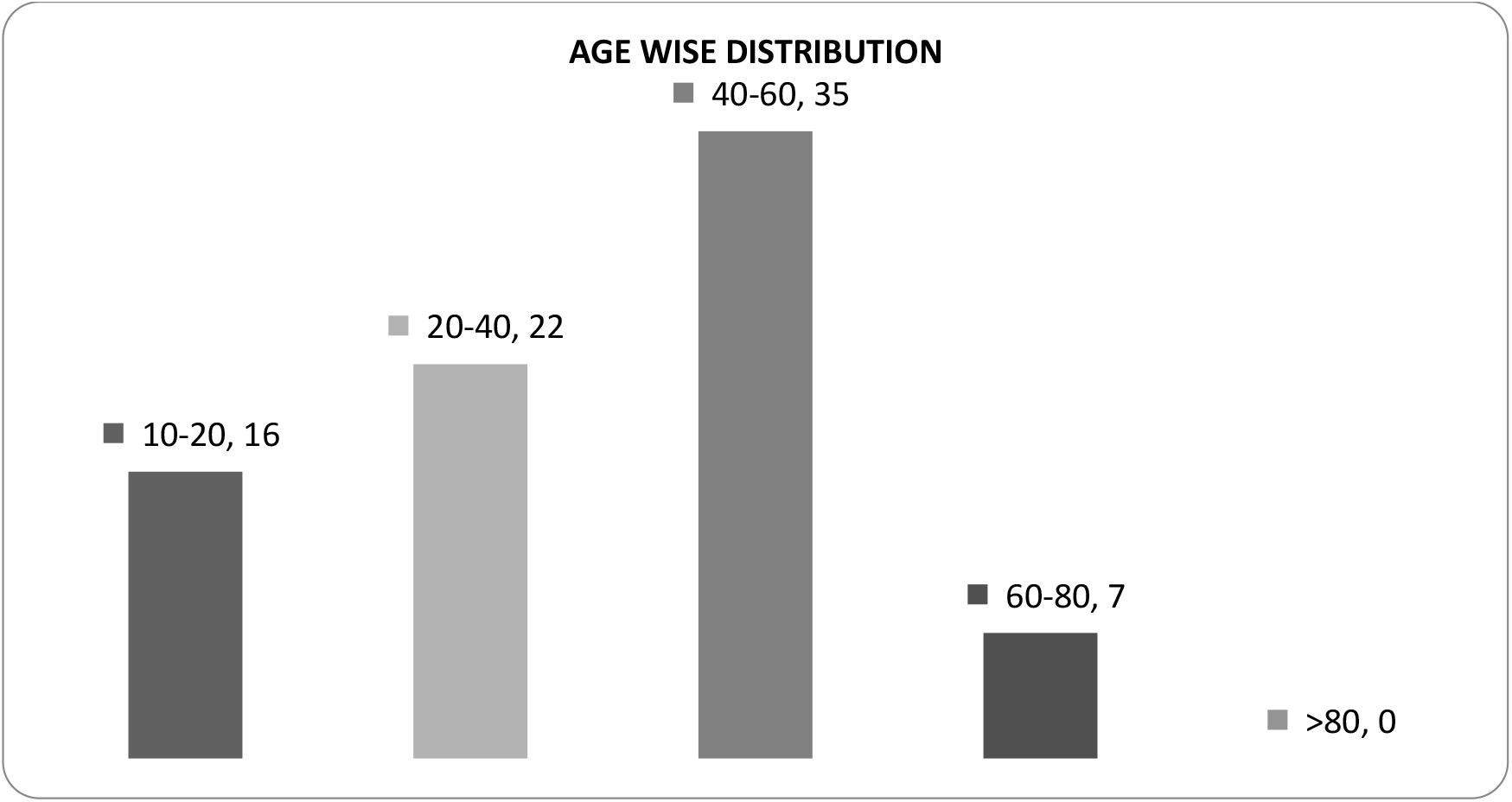

Most common age group affected were 40-60 years of age of 35 numbers, followed by 20-40 years of 22 numbers, followed by 10-20 years of age of 16 numbers, followed by 60-80 years of age of 7 numbers.

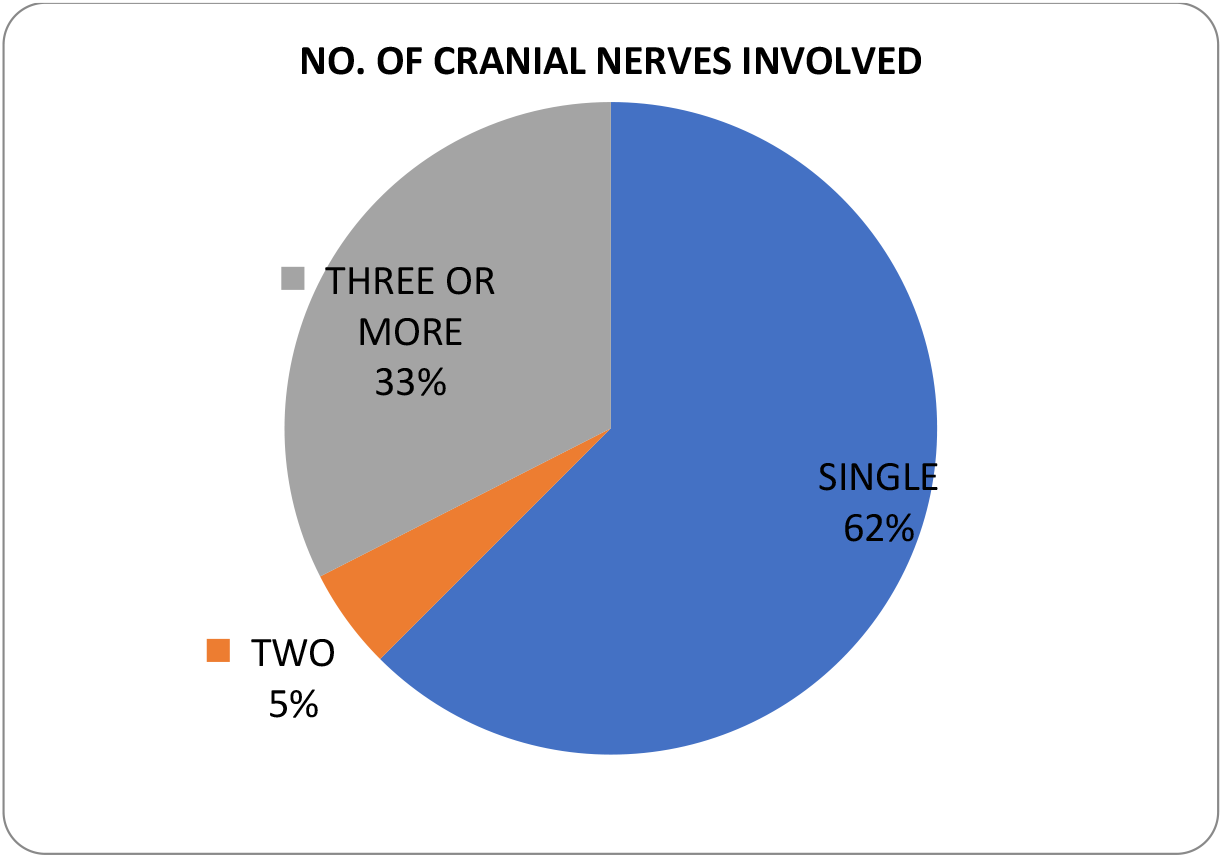

Most common pattern of involvement was the single cranial nerve of 62 % (50); followed by multiple cranial nerves of more than two 33% (26); followed by two cranial nerves 5% (4)

#### B. Cranial Nerve Distribution

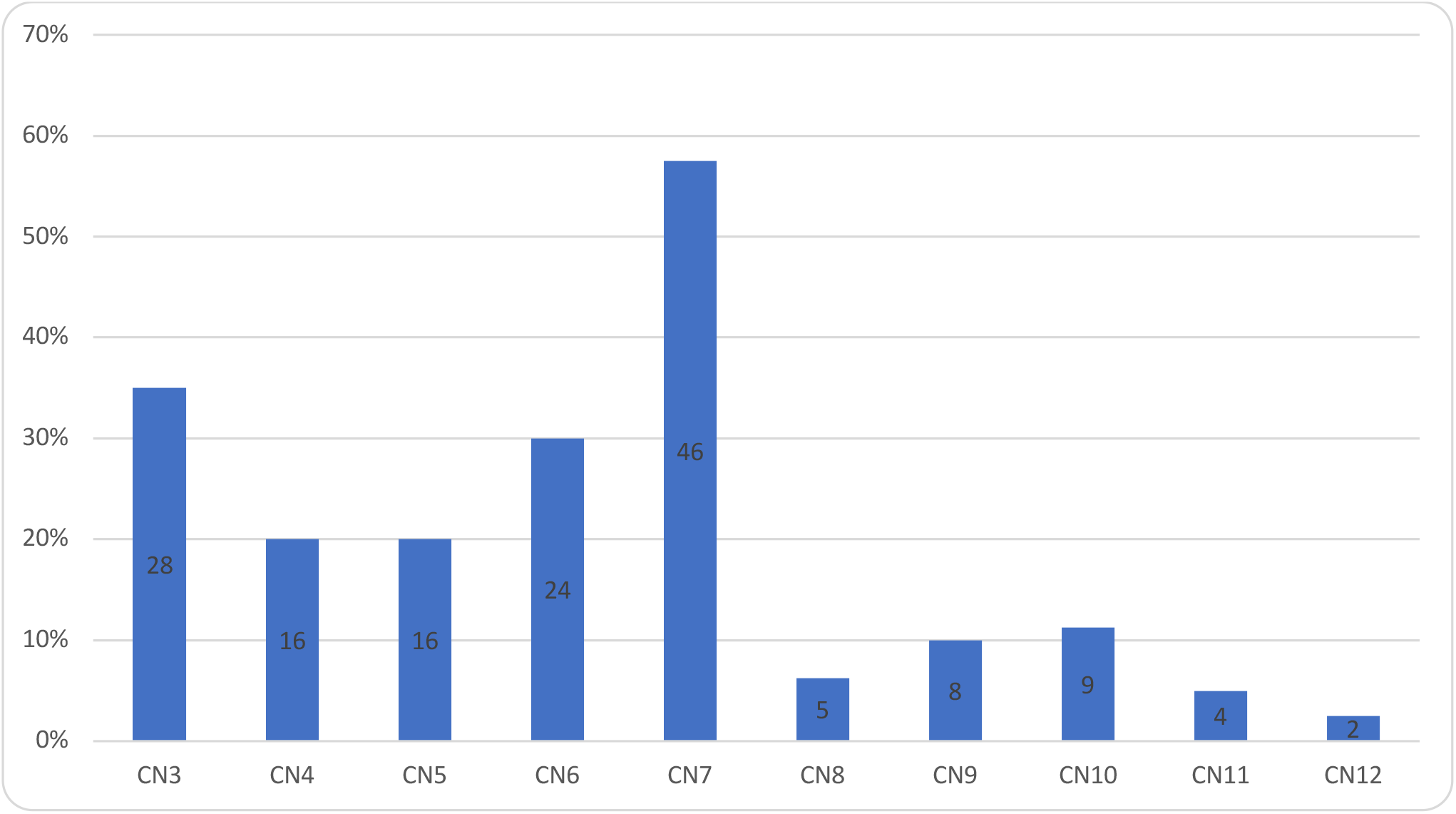

Most common cranial nerve involved was 7^th^ of 58%(46); followed 3^rd^ of 36% (28); 6^th^ of 30% (24); 4^th^ and 5^th^ together of 20 % each (16); followed by 10^th^ of 12 % (9); 9^th^ of 10%(8); 8^th^ of 6%(5); 11^th^ of 5%(4); 12^th^ of 1% (2);

#### C. Intra Axial / Extra Axial

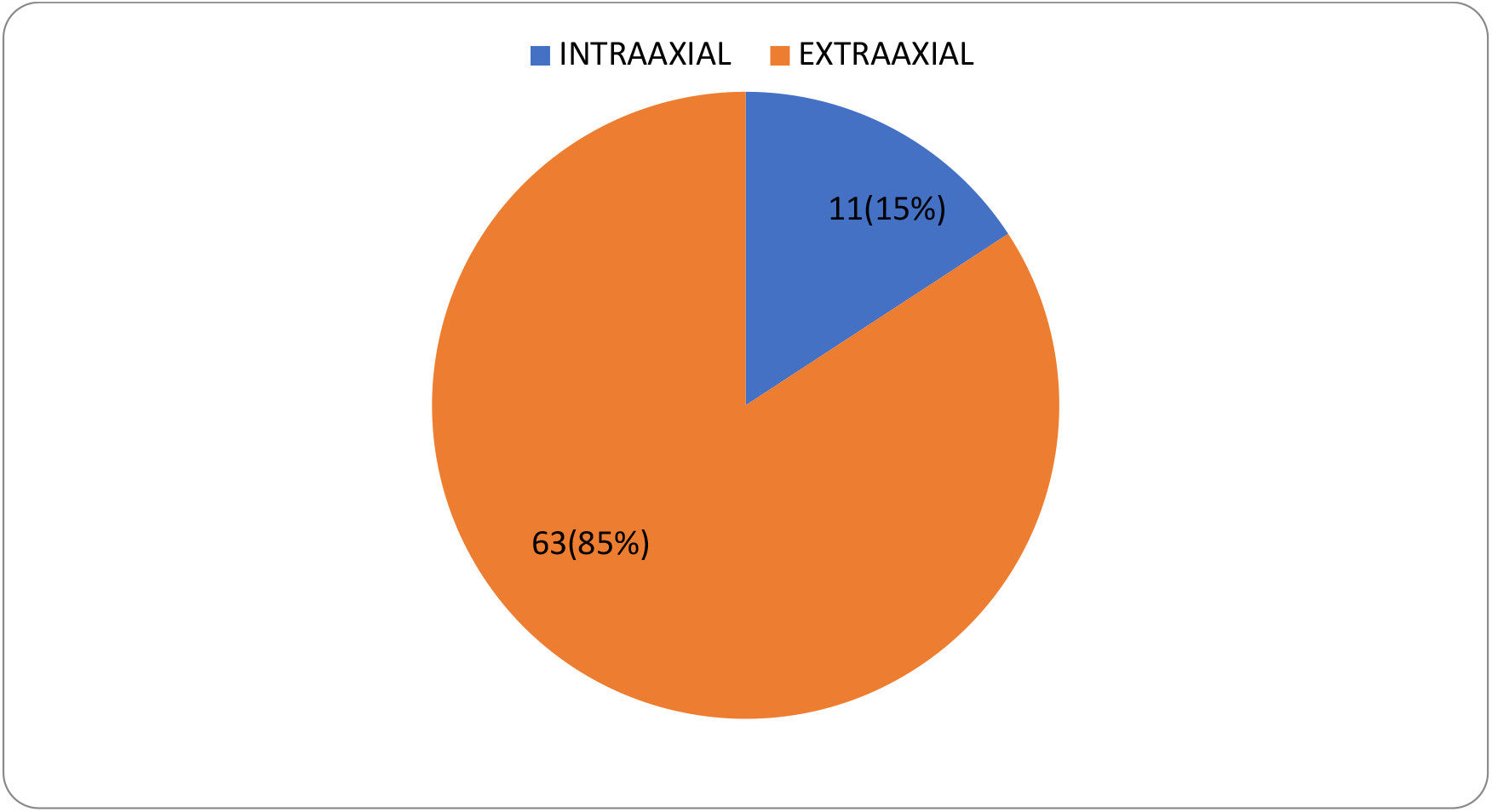

Most common localization was extra-axial of 85 % (63); followed by intra axial of 15% (11)

#### D. Most Common Etiology

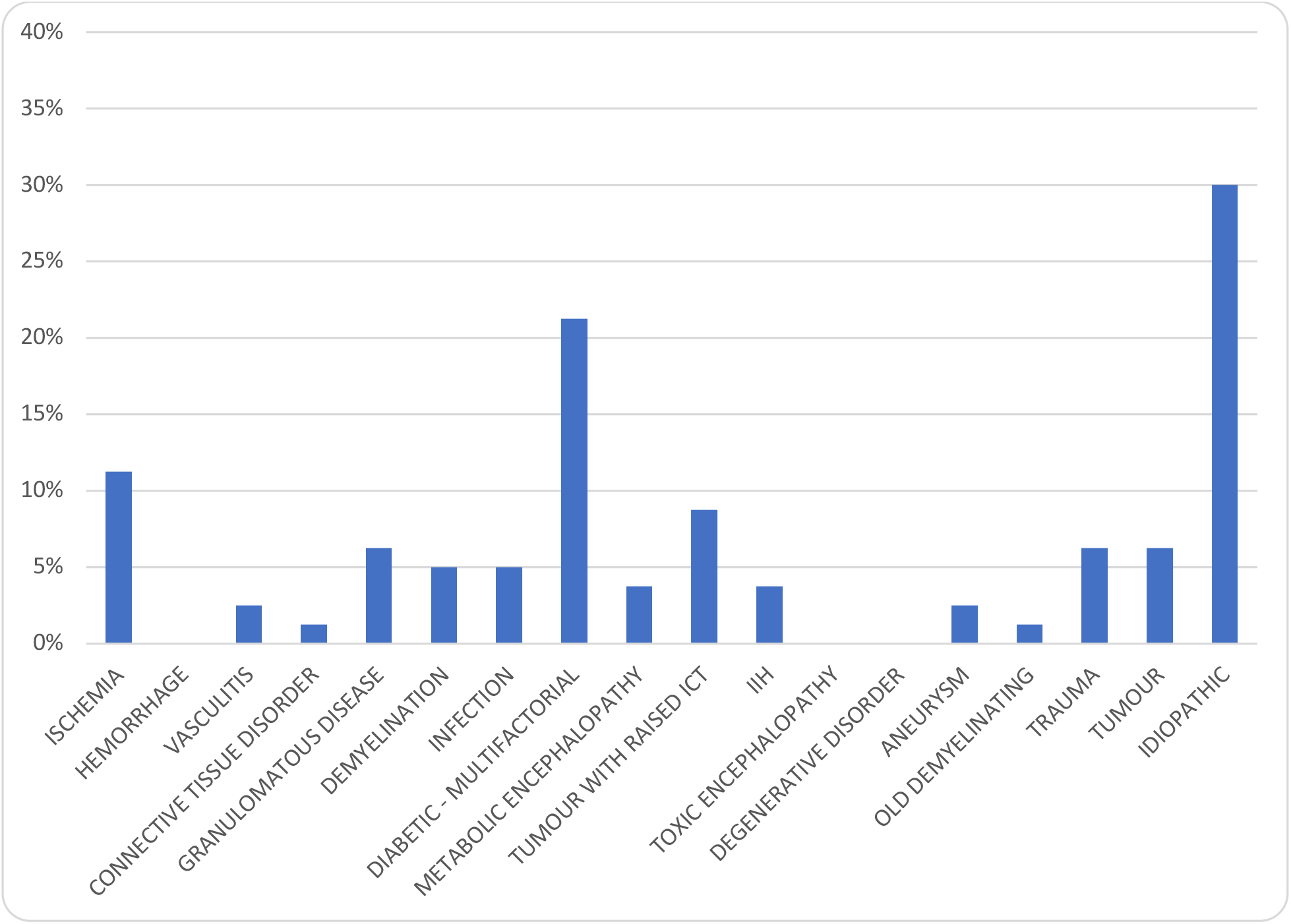

Most common etiology was idiopathic with 30 % (24); followed by diabetes with its multifactorial mechanisms 21 % (17); followed by ischemia 11 % (9); followed by tumor with raised ict of 9 % (7); trauma, tumor without raised ict, granulomatous diseases with 7% of 5 numbers each; followed by demyelination, infection of 6% (4); followed by metabolic encephalopathy, IIH of 5% (3); followed by vasculitis, aneurysm of 4% (2); followed by connective tissue disorder of 2 % (1);

#### E. Risk Factors

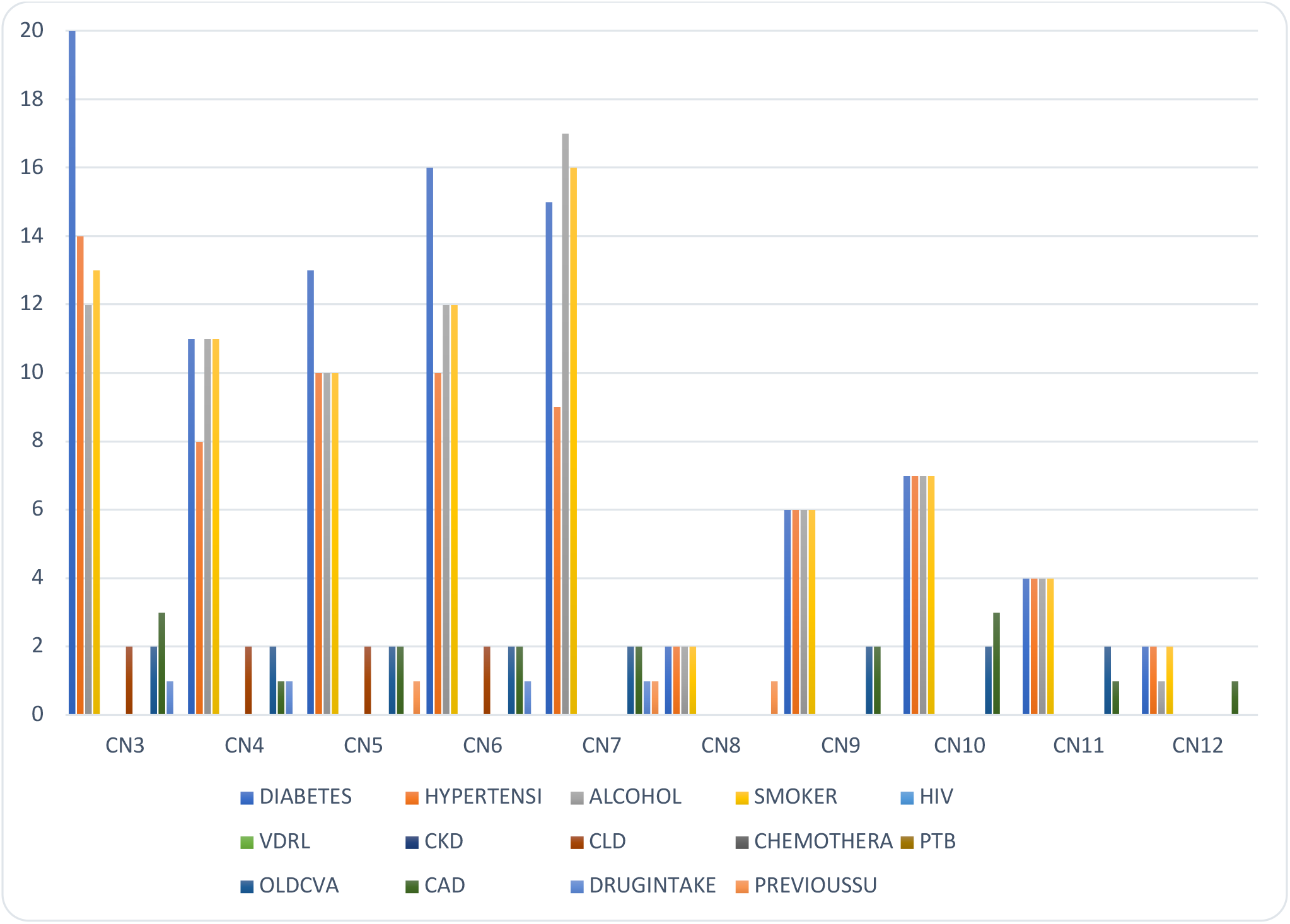

|  | DIABETES | HYPERTENSION | ALCOHOL | SMOKER | HIV | VDRL | CKD | CLD | CHEMOTHERAPY | PTB | OLDCVA | CAD | DRUGINTAKE | PREVIOUS |
| --- | --- | --- | --- | --- | --- | --- | --- | --- | --- | --- | --- | --- | --- | --- |
| CN3 | 20 | 14 | 12 | 13 | 0 | 0 | 0 | 2 | 0 | 0 | 2 | 3 | 1 | 0 |
| CN4 | 11 | 8 | 11 | 11 | 0 | 0 | 0 | 2 | 0 | 0 | 2 | 1 | 1 | 0 |
| CN5 | 13 | 10 | 10 | 10 | 0 | 0 | 0 | 2 | 0 | 0 | 2 | 2 | 0 | 1 |
| CN6 | 16 | 10 | 12 | 12 | 0 | 0 | 0 | 2 | 0 | 0 | 2 | 2 | 1 | 0 |
| CN7 | 15 | 9 | 17 | 16 | 0 | 0 | 0 | 0 | 0 | 0 | 2 | 2 | 1 | 1 |
| CN8 | 2 | 2 | 2 | 2 | 0 | 0 | 0 | 0 | 0 | 0 | 0 | 0 | 0 | 1 |
| CN9 | 6 | 6 | 6 | 6 | 0 | 0 | 0 | 0 | 0 | 0 | 2 | 2 | 0 | 0 |
| CN10 | 7 | 7 | 7 | 7 | 0 | 0 | 0 | 0 | 0 | 0 | 2 | 3 | 0 | 0 |
| CN11 | 4 | 4 | 4 | 4 | 0 | 0 | 0 | 0 | 0 | 0 | 2 | 1 | 0 | 0 |
| CN12 | 2 | 2 | 1 | 2 | 0 | 0 | 0 | 0 | 0 | 0 | 0 | 1 | 0 | 0 |

### 2. AMONG INDIVIDUAL CRANIAL NERVES

#### A. Age Group Distribution

2. AMONG INDIVIDUAL CRANIAL NERVES: A. Age Group Distribution:
| CN | 1-20 | 21-40 | 41-60 | 61-80 | Grand Total |
| --- | --- | --- | --- | --- | --- |
| CN 3 | 2 | 7 | 15 | 4 | 28 |
| CN 4 | 2 | 2 | 9 | 3 | 16 |
| CN 5 | 1 | 2 | 10 | 3 | 16 |
| CN 6 | 4 | 3 | 14 | 3 | 24 |
| CN 7 | 15 | 16 | 13 | 2 | 46 |
| CN 8 | 1 | 2 | 2 | - | 5 |
| CN 9 | 1 | 1 | 3 | 3 | 8 |
| CN 10 | 1 | 1 | 4 | 3 | 9 |
| CN 11 | - | - | 1 | 3 | 4 |
| CN 12 | - | - | - | 2 | 2 |

Overall, the most common age group distribution was around 41-60 years of age; except in cn 7 where 21-49 years were most commonly affected; followed by most common age group distribution was 21-40 years; exception in cn 7 where 1-20 years were the most common group; followed by 41-60 years of age. overall 61-80 years were next in order followed by 1-20 years Males were most commonly affected in all the cranial nerves.

#### B. Number Of Nerves Involved

B. Number Of Nerves Involved:
| CN | SINGLE/TWO/MULTIPLE | Total |
| --- | --- | --- |
| CN 3 | MULTIPLE | 16 |
|  | SINGLE | 11 |
|  | TWO | 1 |
|  | TOTAL | 28 |
| CN 4 | MULTIPLE | 16 |
|  | TOTAL | 16 |
| CN 5 | MULTIPLE | 16 |
|  | TOTAL | 16 |
| CN 6 | MULTIPLE | 19 |
|  | SINGLE | 4 |
|  | TWO | 1 |
|  | TOTAL | 24 |
| CN 7 | MULTIPLE | 13 |
|  | SINGLE | 32 |
|  | TWO | 1 |
|  | TOTAL | 46 |
| CN 8 | MULTIPLE<br>TWO | 4<br>1 |
|  | TOTAL | 5 |
| CN 9 | MULTIPLE<br>TWO | 7<br>1 |
|  | TOTAL | 8 |
| CN 10 | MULTIPLE<br>SINGLE<br>TWO | 7<br>1<br>1 |
|  | TOTAL | 9 |
| CN 11 | MULTIPLE | 4 |
|  | TOTAL | 4 |
| CN 12 | MULTIPLE<br>SINGLE | 1<br>1 |
|  | TOTAL | 2 |

Among the number of nerves involved, 3^rd^, 6^th^,8^th^,9^th^,10^th^ cranial nerve was most commonly presented in multiple combinations followed by single; cn 4^th^, 5^th^, 11^th^, presented only in multiple combinations pattern, 12^th^ both in single as well as multiple combo pattern. whereas, 7^th^ cn was mostly presented in isolated fashion followed by multiple combination pattern;

#### C. ETIOLOGY

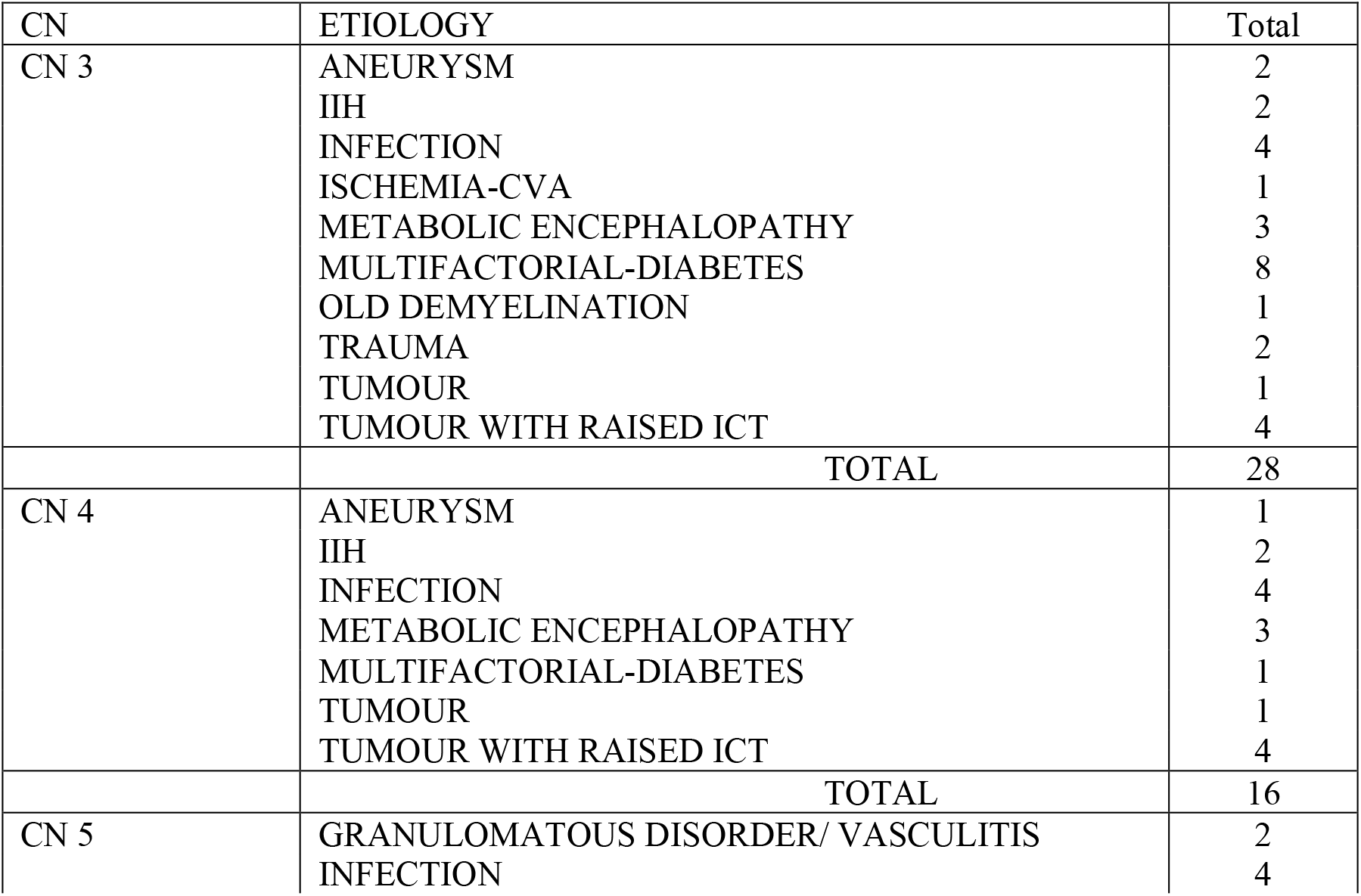
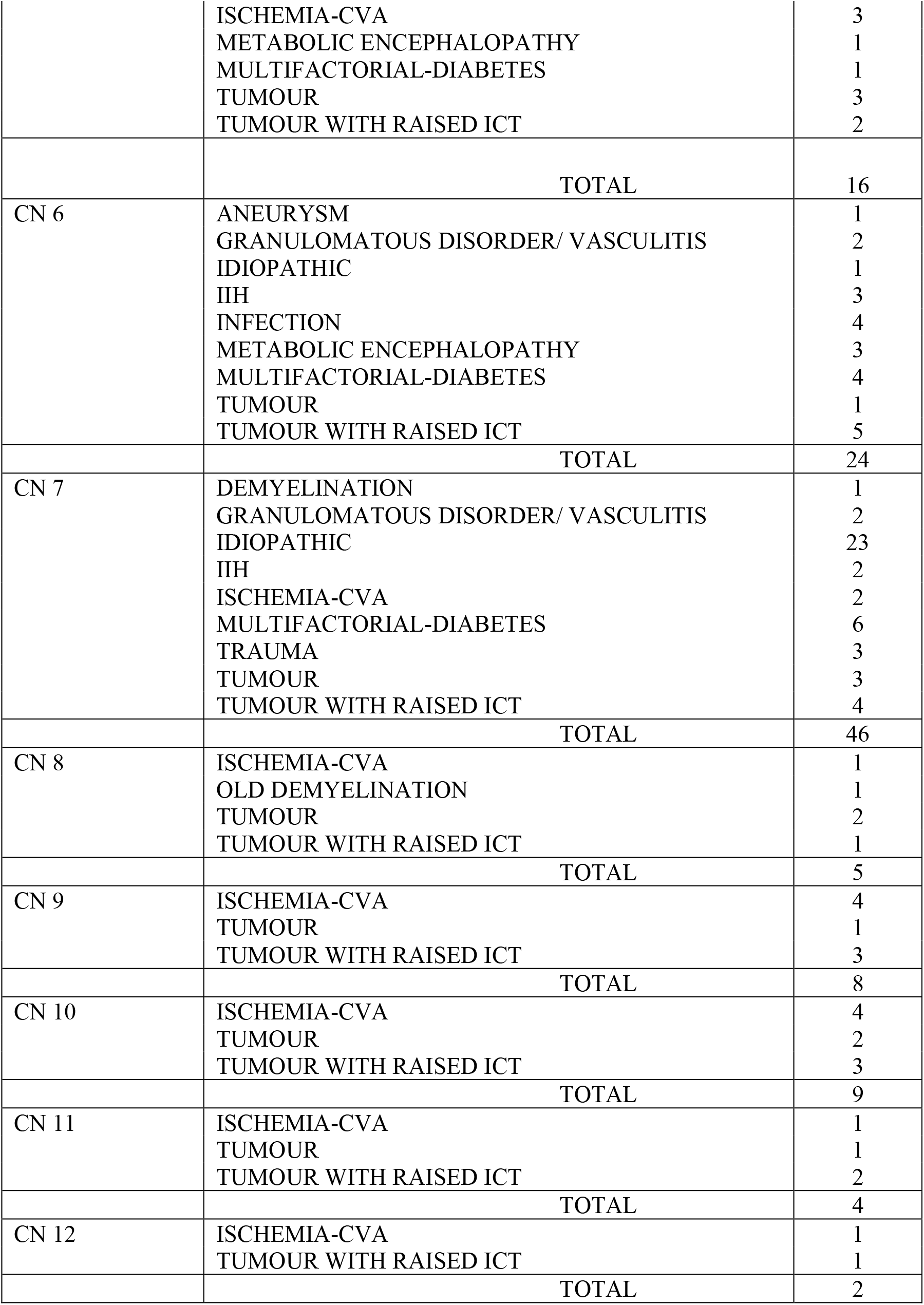

In the 3^rd^ cranial nerve, the most common pathology was due to diabetes; followed by a cavernous sinus infection and tumor with raised ict, followed by Wernicke encephalopathy; then comes trauma, aneurysm, iih.

In the 4^th^ cranial nerve; the most common pathology was cavernous sinus infection followed by tumor with raised ict; followed by metabolic encephalopathy, then comes diabetes; aneurysm

In the 5^th^ cranial nerve-the most common pathology was cavernous sinus infection; followed by post circulation stroke; followed by tumor with raised ict

In the 6^th^ cranial nerve; the most common pathology was tumor with raised ict, cavernous sinus infection; diabetes, followed by iih, metabolic encephalopathy, followed by vasculitis, aneurysm In the 7^th^ cranial nerve, the most common pathology was idiopathic; followed by diabetes, followed by trauma, tumor, followed by iih, post circulation strokes, and vasculitis

In the 8^th^ cranial nerve; the most common pathology was tumor-cp angle; followed by stroke and demyelination

In the 9^th,^ 10^th^ cranial nerve, the most common pathology was due to ischemia – posterior circulation strokes, followed by tumor

In the 11^th^ cranial nerve, the most common pathology was tumor followed by ischemia; In the 12^th^ cranial nerve, both ischemia and tumor were equally involved

### D. INTRA AXIAL/ EXTRA AXIAL

**D. INTRA AXIAL/ EXTRA AXIAL:**
| CN | INTRAAXIAL/EXTRAAXIAL | Total |
| --- | --- | --- |
| CN 3 | EXTRAAXIAL | 23 |
|  | INTRAAXIAL | 5 |
|  | TOTAL | 28 |
| CN 4 | EXTRAAXIAL | 13 |
|  | INTRAAXIAL | 3 |
|  | TOTAL | 16 |
| CN 5 | EXTRAAXIAL | 12 |
|  | INTRAAXIAL | 4 |
|  | TOTAL | 16 |
| CN 6 | EXTRAAXIAL | 21 |
|  | INTRAAXIAL | 3 |
|  | TOTAL | 24 |
| CN 7 | EXTRAAXIAL | 44 |
|  | INTRAAXIAL | 2 |
|  | TOTAL | 46 |
| CN 8 | EXTRAAXIAL | 3 |
|  | INTRAAXIAL | 2 |
|  | TOTAL | 5 |
| CN 9 | EXTRAAXIAL | 4 |
|  | INTRAAXIAL | 4 |
|  | TOTAL | 8 |
| CN 10 | EXTRAAXIAL | 5 |
|  | INTRAAXIAL | 4 |
|  | TOTAL | 9 |
| CN 11 | EXTRAAXIAL | 3 |
|  | INTRAAXIAL | 1 |
|  | TOTAL | 4 |
| CN 12 | EXTRAAXIAL | 1 |
|  | INTRAAXIAL | 1 |
|  | TOTAL | 2 |

Most Common Localisation For All The Cranial Nerves Was Extraaxial Lesion Followed By Intraaxial; Where As In 9^th^ And 12^th^ Cranial Nerves, There Were Equal Distribution Of Pathology

## DISCUSSION

Maximum of the preceding literature concerning single/more than one cranial nerve palsies is either from the western nations or is in the form of case collection and case reviews [3, 8]. The etiological spectrum of more than one cranial nerve palsies is probably exclusive in a developing country like India.

We observed that the seventh cranial nerve is the isolated most common cranial nerve involved, followed by 3rd and 6th then coming 4th, fifth cranial nerves observed by others.

Idiopathic-stylomastoid foramen, superior orbital fissure, cavernous sinus, orbital apex, the base of the skull have been the order of anatomical localization within the extra-axial involvement of cranial nerves. Posterior circulation strokes followed by brainstem demyelination have been the maximum commonplace causes and localization within the intra axial involvement of cranial nerves. A number of the multiple cranial nerve combination involvement, the biggest collection of more than one cranial nerve palsies was published with the aid of Keane [3]. within Keane’s series, the maximum common cranial nerve involved was also the abducens (VI) and the most common site become cavernous sinus. but the study through Keane become retrospective and included patients with neuromuscular junction disorders, brainstem syndromes, and cranial nerve palsies as a part of generalized neuropathic procedures like Guillain-Barre syndrome [3].

Multifactorial reason by way of diabetes was the most common etiology in our study, in contrast to western literature where tumor became reported to be the maximum common reason for multiple cranial neuropathies. This becomes expected, as the prevalence of diabetes is lots better in growing countries like India, in comparison to the evolved ones, followed by Hypertension, Alcoholism and Smoking, Ischemic Etiology for all the Cranial Nerves. Similar findings had been highlighted via Bhatkar and co-workers [9]. Tuberculous and fungal infections were the maximum not unusual infections encountered in our study. Tuberculous meningitis is a not unusual purpose of a couple of cranial nerve palsy in India. The examination by way of Sharma and co-employees observed that cranial nerve involvement became found in 38% instances of tuberculous meningitis, with about 10% having more than one cranial nerve involvement [5]. The fungal infections encountered in our examination generally offered with orbital apex, superior fissure, and cavernous syndromes; mucormycosis became the most common fungal infection determined in our study. Bhatkar et al. stated 24.6% cases of fungal contamination, aspergillosis being the most common cause [9]. Bilateral involvement and speedy onset vision loss have been seen more in fungal infection compared to other aetiologies, additionally proven by way of Chua et al [10]. Imaging findings in patients with fungal infection showed para-nasal sinusitis, multiple focal regions of bony destruction, heterogeneous signal intensity and contrast enhancement of orbit, nasal sinuses, and cavernous sinus. several studies were cited in the literature concerning radiological findings in CNS fungal infections [11, 12].

Tolosa Hunt syndrome, diagnosed by way of ICHD-III standards [7] was seen in eleven% of our cases. This frequency became decrease compared to the previous case series [3,9]. Tolosa Hunt syndrome is an essential cause of painful ophthalmoplegia, however, the etiology remained unknown [13]. In our study, all six patients had MRI abnormalities, like those mentioned in advance [14, 15] strong gadolinium enhancement of cavernous sinus wall with or without focal narrowing of ICA inside the cavernous sinus. Two out of six sufferers had kind 2 diabetes mellitus. Such co-lifestyles have been mentioned in current case reviews [16].

Metastasis observed with the aid of schwannoma was the maximum not unusual tumors (benign/malignant) main to more than one cranial neuropathies in our study. in Keane’s series,[3] schwannoma become discovered to be the most common tumor, whilst inside the cavernous sinus syndrome series of Bhatkar et al., [9] Metastasis changed into more common than the primary tumor.

For some of the lower cranial nerves, the most common combination becomes the IX, X, and XII cranial nerves. Out of the six cases, 3 had been of tumors (one case every of pontine glioma, jugular schwannoma, and nerve sheath tumor extending from craniovertebral junction to C2 vertebra). Common reasons for lower cranial nerve palsies include vascular, stressful, neoplastic, iatrogenic, and infective [17]. Additionally, trauma, vasculitis, which is a commonplace group of problems supplying with multiple cranial neuropathies, was also encountered in our study. A bigger study would substantiate all the various companies of issues that can be pronounced in literature.

Therefore, to finish, amongst more than one cranial neuropathies, cavernous sinus syndrome and orbital apex syndrome have been the most common anatomical pattern of involvement, and infections which include tuberculous and fungal were the most common reasons for cranial nerve palsy in our collection. Bigger studies with long-term follow-up are needed in India to evaluate the causal association. despite exhaustive workup, an etiological prognosis can’t be reached in each case [18-20].

## Data Availability

Data will be shared on contacting the authors

